# Comparison of COVID-19 case and death counts in the United States reported by four online trackers: January 22-May 31, 2020

**DOI:** 10.1101/2020.06.20.20135764

**Authors:** Jinkinson Smith

## Abstract

The first purpose of this study is to describe a project focused on comparing the numbers of COVID-19 cases and deaths in the United States reported by four different online trackers, namely, those maintained by USAFacts, the New York Times, Johns Hopkins University, and the COVID Tracking Project. The second purpose of this study is to present results from the first five months of 2020 (January 22-May 31, 2020). This project is ongoing, so it will be updated regularly as new data from each of these trackers become available. Based on the time period included, the NYT has reported more cases than any of the other three trackers since late March/early April, and COVID Tracking Project has reported fewer deaths than any of the other three trackers since mid-March. It is hoped that the discrepancies identified by this project will provide avenues for research on their causes.

## Introduction

This study aims to describe a regularly updated project I have been (and still am) conducting, the aim of which is to compare the number of COVID-19 cases and deaths in the United States reported by four different online trackers. In doing this, I hope to provide evidence either for or against the hypothesis that there are systematic differences between the values reported by the different trackers.

The four COVID-19 United States-specific datasets I will be comparing are from USAFacts, the New York Times (hereafter NYT), Johns Hopkins University (hereafter JHU), and the COVID Tracking Project. First of all, there are some differences in the start dates for each of the four datasets. All of them started on January 22, except for the NYT, which started a day earlier (January 21).

The total number of cases in the United States reported by each tracker were compared over time for each date from the first date including all four trackers (i.e. January 22, 2020) to the last day of May (i.e. May 31, 2020). This comparison was done to shed light on the extent to which the number of cases reported by the four included trackers, namely COVID Tracking Project, NYT, JHU, and USAFacts (hereafter simply “the four trackers”) differed and how these differences had changed over time. Below, I give the URLs from which I obtained the data from each source used in this study.

1. The COVID Tracking Project data was obtained from this link: https://covidtracking.com/data/us-daily/
2. The NYT data was obtained from this link: https://github.com/nytimes/covid-19-data/blob/master/us.csv
3. The JHU data was obtained from this link: https://github.com/CSSEGISandData/COVID-19/tree/master/csse_covid_19_data/csse_covid_19_daily_reports_us
4. Finally, the USAFacts data was obtained from this link for cases: https://usafactsstatic.blob.core.windows.net/public/data/covid-19/covid_confirmed_usafacts.csv and this link for deaths: https://usafactsstatic.blob.core.windows.net/public/data/covid-19/covid_deaths_usafacts.csv

## Results

### Comparing the number of cases across the four trackers

The total number of cases over time reported by each of the four trackers is shown graphically in Figure 1. It is clear that all four match up very closely, as would be expected if they are both largely successful in their shared goal of measuring the same underlying value.

**Figure 1:**
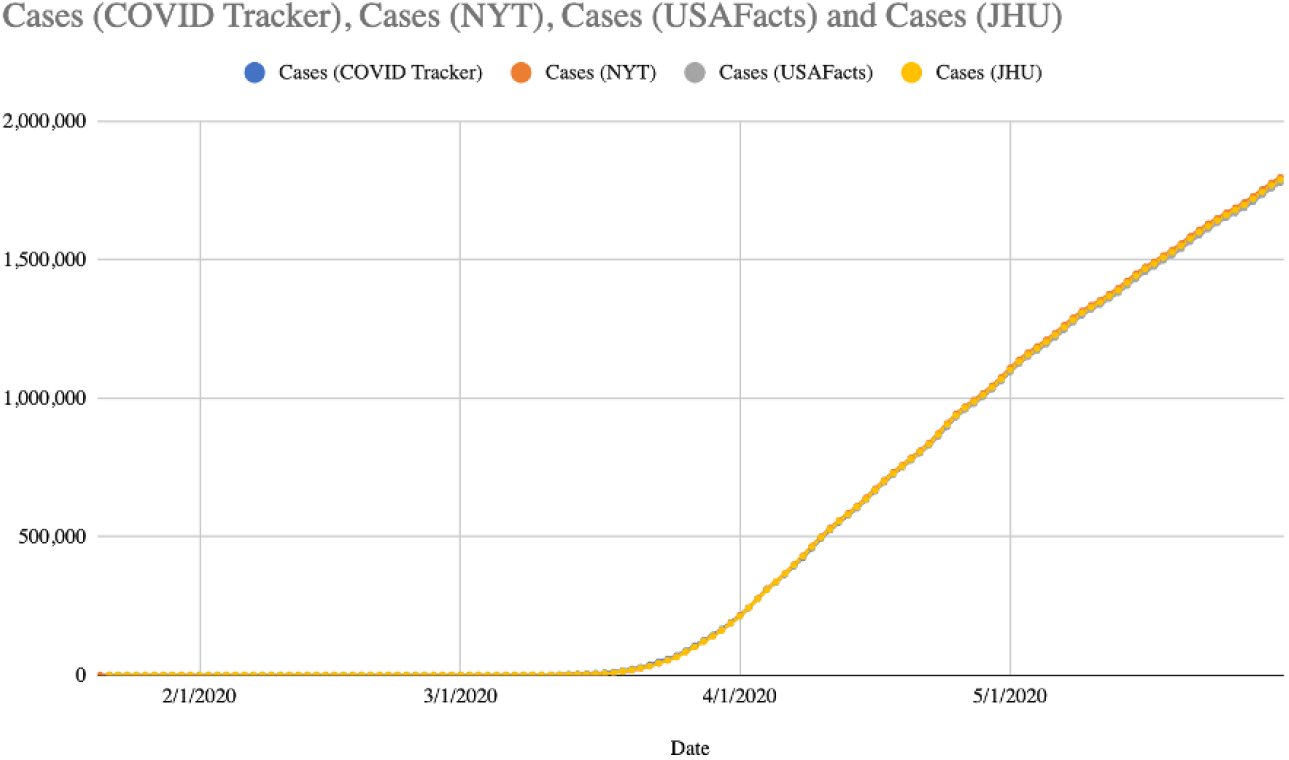
Total number of cases of COVID-19 reported in the US according to four different trackers

The absolute difference between the numbers of total cases reported by each possible combination of the four trackers is shown in Fig. 2 below. (By “each possible combination” I mean one of the six following: NYT and COVID Tracking Project, NYT and USAFacts, NYT and JHU, COVID Tracking Project and USAFacts, COVID Tracking Project and JHU, and USAFacts and JHU.) Each of the six lines corresponds to the result of subtracting the number of cases reported by the second tracker in the legend for that line from the number reported by the first tracker on a given day. For example, on March 10, the line corresponding to “NYT-USAFacts” has a value of −21, because on that date, NYT reported 1,018 cases and USAFacts reported 1,039, so NYT-USAFacts = 1,018-1,039 = −21.

**Figure 2:**
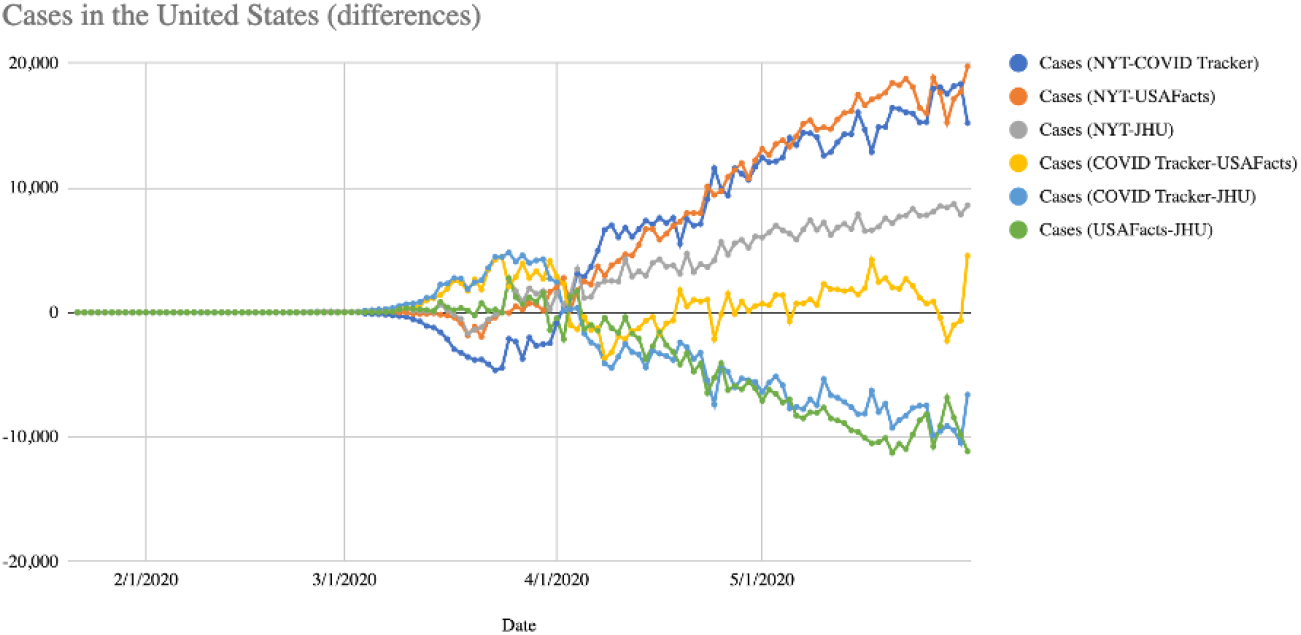
Difference in total number of COVID-19 cases reported in the United States between four different trackers

From the above Fig. 2 it is clear that NYT is now, and for some time has been, reporting noticeably more cases nationally compared to either COVID Tracking Project or USAFacts. The absolute difference in reported cases between NYT and COVID Tracking Project, as well as that between NYT and USAFacts, has also been increasing for most of the past two months (i.e. all of April and May) Another pattern that has been ongoing for some time now is that NYT is reporting more cases than JHU, and the difference between these two has also generally been increasing over the past two months. Nevertheless, it is noteworthy that JHU is reporting more cases than either COVID Tracking Project or JHU, and that, again, both the gap between JHU and COVID Tracking Project and the gap between USAFacts have been steadily increasing for most of the last two months. Finally, COVID Tracking Project and USAFacts have been reporting the most similar numbers of nationwide cases of any two trackers included here, as is evident from the differences line for COVID Tracking Project-USAFacts in Fig. 2 having mostly fluctuated closer to the x-axis than any of the other five.

### COVID Tracking Project vs. JHU

During all of March and the first few days of April, COVID Tracking Project reported more cases nationwide than did JHU. (Note that all subsequent figures refer to national data unless indicated otherwise.) However, on April 5, this changed and JHU’s case count surpassed that of COVID Tracking Project. This has remained the case ever since, so that as of May 31, JHU was reporting 6,602 more cases nationally than COVID Tracking Project.

The Pearson correlation between the number of cases reported by COVID Tracking Project and JHU was 0.9999971.

### COVID Tracking Project vs. NYT

On March 3, 2020, COVID Tracking Project was reporting just 1 more case than NYT. COVID Tracking Project’s lead over NYT then grew until it reached its peak of 4,643 cases on March 23, at which point it started shrinking. By April 2, this lead had been completely eliminated and NYT was now on top, reporting 467 more cases than COVID Tracking Project. Since then, the NYT’s lead over COVID Tracking Project has grown to 15,194 cases (as of May 31).

The Pearson correlation between the number of cases reported by COVID Tracking Project and NYT was 0.9999970.

### COVID Tracking Project vs. USAFacts

From January 24 to February 29 inclusive, USAFacts was reporting more cases nationally than COVID Tracking Project. But this changed on March 1, when COVID Tracking Project reported a total of 4 more cases than USAFacts. This was the first time that COVID Tracking Project reported more cases than USAFacts, a pattern which continued until April 3, when the number of USAFacts cases surpassed the number reported by COVID Tracking Project by 1,017 (by contrast, the day before, April 2, COVID Tracking Project was reporting 2,291 more cases than USAFacts). Since then, these two trackers have exchanged positions relative to one another ten times (as of May 31).

The Pearson correlation between the number of cases reported by COVID Tracking Project and USAFacts was 0.9999970.

### NYT vs. USAFacts

Every day from March 26 to May 31 inclusive, NYT has reported more cases than USAFacts. On March 26, NYT reported 495 more cases than USAFacts, and by May 31, this lead had grown to 19,736 cases.

The Pearson correlation between the number of cases reported by NYT and USAFacts was 0.9999990.

### NYT vs. JHU

Every day from March 25 to May 31 inclusive, NYT reported more cases than JHU. During this time, the lead NYT’s case count has over that of JHU has grown from 2,686 on March 25 to 8,592 on May 31.

The Pearson correlation between the number of cases reported by NYT and JHU was 0.9999996.

### USAFacts vs. JHU

Every day from April 5 to May 31 inclusive, JHU has reported more cases nationally compared to USAFacts. On April 5, JHU reported 1,690 more cases nationally than did USAFacts, and this value grew to 6,602 more than USAFacts by May 31.

The Pearson correlation between the number of cases reported by USAFacts and JHU was 0.9999987.

### Comparing the number of deaths across the four trackers

Fig. 3 below shows the total number of COVID-19 deaths in the United States reported by the four trackers. As is evident from this figure, COVID Tracking Project (blue line) has reported significantly fewer deaths than any of the other three trackers (orange, gray, and yellow) for some time. The other three trackers’ death counts have tended to cluster more closely together.

**Figure 3:**
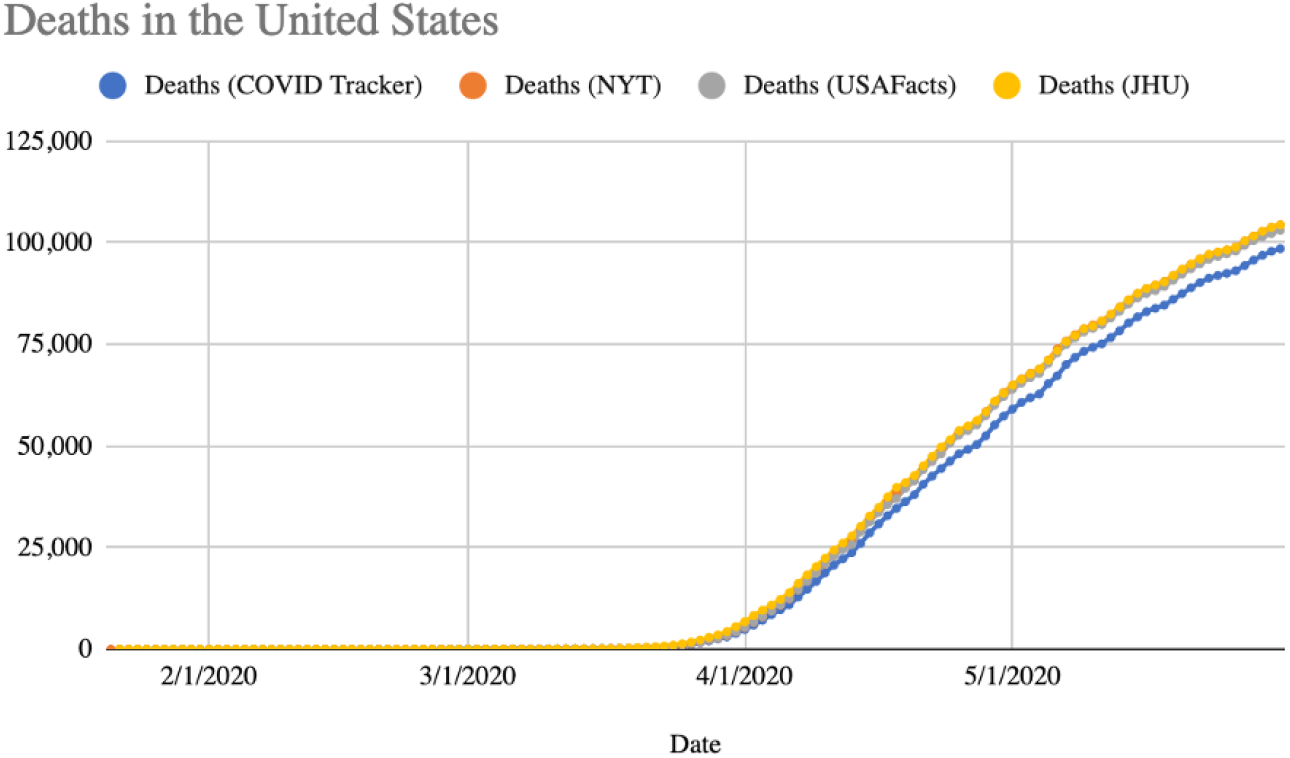
Total number of deaths from COVID-19 reported in the US according to each of the four trackers

The differences between the four trackers in terms of the number of total deaths they reported over time is shown in Fig. 4; this is analogous to Fig. 2 mentioned earlier, only that one applied to cases rather than to deaths. This figure clearly shows that NYT has been reporting more deaths than COVID Tracking Project since late March, and that the number of excess deaths reported by NYT compared to COVID Tracking Project has been fairly steady throughout the month of May. The grey line (NYT-JHU) has been very close to the x-axis for the entire month of May, indicating that NYT and JHU have been the two closest during this month in terms of the total number of deaths they have reported. For roughly the second half of April and all of May, we also see the NYT reporting more deaths than USAFacts, but only slightly more, not nearly so many more as NYT is reporting compared to COVID Tracking Project.

**Figure 4:**
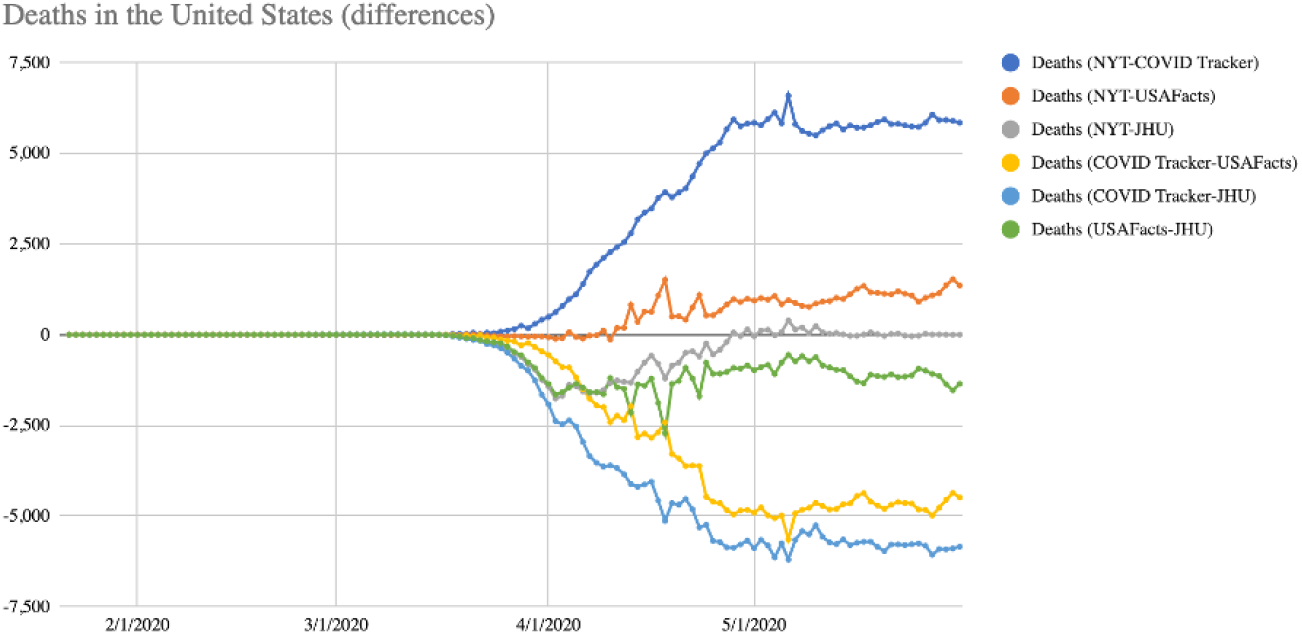
Differences between the total number of COVID-19 deaths in the United States reported by each of four trackers

### COVID Tracking Project vs. JHU

As of May 31, there were 5,845 more deaths reported by JHU than by COVID Tracking Project. The last time JHU’s death toll was not higher than that of COVID Tracking Project was March 15, when the former exceeded the latter by just 6 deaths. March 29 was the last time JHU’s death toll exceeded that of COVID Tracking Project by less than 1,000 deaths (in that case, the difference between the two was 991 deaths).

The Pearson correlation between the number of deaths reported by COVID Tracking Project and JHU was 0.9996166.

### COVID Tracking Project vs. NYT

As of May 31, there were 5,845 more deaths in the US reported by the NYT than by COVID Tracking Project, and the last time that COVID Tracking Project reported more deaths nationally than NYT (as well as the last time NYT did not report more deaths than COVID Tracking Project) was over two months earlier on March 17, when COVID Tracking Project reported 3 more deaths nationally than NYT. Every day since (and including) April 5, the NYT’s death count has exceeded that of COVID Tracking Project by at least 1,000 (meaning April 4, when NYT’s death count exceeded COVID Tracking Project’s by 979 deaths, is the last time that NYT’s lead over COVID Tracking Project in this regard was less than 1,000 deaths). The last date when the NYT did not report at least 5,000 more deaths nationally than did COVID Tracking Project was May 11, when NYT reported 4,980 more deaths than COVID Tracking Project. Every day from May 12 to May 31 inclusive, the NYT has reported between 5,000 and 6,000 more deaths than COVID Tracking Project.

The Pearson correlation between the number of deaths reported by COVID Tracking Project and NYT was 0.9997858.

### COVID Tracking Project vs. USAFacts

As of May 31, there were 4,489 more deaths in the US reported by USAFacts than by COVID Tracking Project, and the last time that USAFacts did not report more deaths nationally than COVID Tracking Project was March 17, when the two trackers reported an identical number of deaths. (The last time COVID Tracking Project reported more deaths than USAFacts was two days earlier, March 15, when it reported 5 more deaths than USAFacts.) As with the NYT (as noted just above), USAFacts’ death count has exceeded that of COVID Tracking Project by over 1,000 every day starting April 5. The last date when this excess was less than 1,000 was April 4, when USAFacts reported 906 more deaths than COVID Tracking Project.

The Pearson correlation between the number of deaths reported by COVID Tracking Project and USAFacts was 0.9998093.

### NYT vs. USAFacts

As of May 31, there were 778 more deaths in the US reported by NYT than by USAFacts, and the last time that NYT did not report more deaths nationally than USAFacts (as well as the last time USAFacts reported more deaths than NYT) was April 10, when USAFacts reported 133 more deaths than NYT.

The Pearson correlation between the number of deaths reported by NYT and USAFacts was 0.9999855.

### NYT vs. JHU

As of May 31, the NYT and JHU trackers were reporting exactly the same number of deaths in the US. These two trackers were relatively close to each other during the month of May: for example, they were less than 100 deaths apart from May 11 to May 31 inclusive, and they have been less than 1,000 deaths apart from April 19 to May 31 inclusive.

The Pearson correlation between the number of deaths reported by NYT and JHU was 0.9999111.

### USAFacts vs. JHU

As of May 31, JHU was reporting 778 more deaths in the US than USAFacts. This was part of a clear long-term pattern: from March 17 to May 31 inclusive, JHU reported more deaths than USAFacts.

The Pearson correlation between the number of deaths reported by USAFacts and JHU was 0.9998943.

## Discussion

This study presents comparisons of the number of total US cases and deaths reported by the NYT, JHU, USAFacts, and COVID Tracking Project. Very strong correlations were consistently found between the values reported by each of the trackers, indicating a high level of agreement. Nevertheless, there were some conspicuous discrepancies between some of the trackers, with the NYT reporting consistently more cases than COVID Tracking Project or USAFacts (and to a lesser extent JHU), beginning in late March/early April. The lead NYT’s reported number of cases has over those reported by COVID Tracking Project and USAFacts has increased steadily since early April, and the magnitude of NYT’s lead over COVID Tracking Project in this regard has consistently been roughly the same as that of NYT’s lead over USAFacts. NYT’s lead in number of reported cases over JHU has also been increasing steadily since early April, but at a significantly slower rate than has NYT’s lead over COVID Tracking Project or USAFacts.

Starting around the same time, NYT began to report significantly more deaths than COVID Tracking Project nationally, and this number continued increasing steadily for about a month thereafter; it has since remained relatively flat. On April 10, NYT began reporting more deaths than USAFacts (though NYT’s lead over USAFacts in the number of reported deaths has since continuously been much smaller than was its lead over COVID Tracking Project in this regard). NYT has reported almost exactly the same number of deaths as JHU since roughly early May. COVID Tracking Project has consistently reported fewer deaths than any of the other three trackers since March 15 or March 17, depending on the tracker. In general, the absolute difference in the number of deaths between any two of the four trackers included here has been relatively stable throughout the month of May.

Further research is warranted as to why that NYT’s tracker is reporting systematically more cases than the other three, especially COVID Tracking Project and USAFacts, as well as why the COVID Tracking Project is reporting systematically fewer deaths than any of the other three trackers.

## Data Availability

I obtained the data itself from one of the four trackers’ freely available websites/repositories. The data I combined into a single spreadsheet is available at my own github at this link: https://github.com/jinkinson/COVID-comparison/blob/master/covid-19%20comparison.csv

